# ROC plot and AUC with binary classifiers: pragmatic analysis of cognitive screening instruments

**DOI:** 10.1101/2021.03.09.21253194

**Authors:** Gashirai K Mbizvo, Andrew J Larner

**Author notes:** Correspondence: AJ Larner, Cognitive Function Clinic, Walton Centre for Neurology and Neurosurgery, Lower Lane, Fazakerley, Liverpool, L9 7LJ, United Kingdom.

## Abstract

Receiver operating characteristic (ROC) plots are a performance graphing method showing the relative trade-off between test benefits (true positive rate) and costs (false positive rate) with the area under the curve (AUC) giving a scalar value of test performance. It has been suggested that ROC and AUC may be potentially misleading when examining binary predictors rather than continuous scales. The purpose of this study was to examine ROC plots and AUC values for two binary classifiers of cognitive status (applause sign, attended with sign), a cognitive screening instrument producing categorical data (Codex), and a continuous scale screening test (Mini-Addenbrooke’s Cognitive Examination), the latter two also analysed with single fixed threshold tests. For each of these plots, AUC was calculated using different methods. The findings indicate that if categorical or continuous measures are dichotomised then the calculated AUC may be an underestimate, thus affecting screening or diagnostic test accuracy which in the context of clinical practice may prove to be misleading.

## Introduction

One of the methods frequently used in the evaluation of screening or diagnostic tests for disease is the construction of a receiver operating characteristic (ROC) curve or plot.^1-3^ This is a graphical representation of the cumulated results of a quantitative test accuracy study across all possible test cut-offs, plotting Sensitivity (Sens) or true positive rate (TPR) on the ordinate against false positive rate (FPR) or 1 – specificity (1 – Spec) on the abscissa.

A measure of how accurately a screening or diagnostic test is able to capture those with and without disease (i.e. its discriminatory ability) may be derived from the area under the ROC curve (AUC).^4,5^ Methods for calculation of AUC are mainly based on a non-parametric statistical test, the Wilcoxon rank-sum test, namely the proportion of all possible pairs of non-diseased and diseased test subjects for which the diseased result is higher than the non-diseased one plus half the proportion of ties.^6^

The performance of a random classifier (i.e. a test which has no discriminatory ability above random chance) is shown by the diagonal line through ROC space (where y = x, or Sens = 1 – Spec, or TPR = FPR) and gives AUC = 0.5. ROC plots ideally approximate the top left hand (“north west”) corner of the ROC space, at coordinates (0,1), where for a perfect classifier AUC = 1. Several qualitative schemata for the classification of AUC values between 0.5 and 1 are available.^7-9^ Symmetrical ROC curves have a constant diagnostic odds ratio (DOR), with DOR = 1 for a random classifier and DOR = ∞ for a perfect classifier. In addition to the rank-sum test method, AUC values may also be calculated based on DOR.^10^

Calculating AUC is very popular in diagnostic accuracy literature because whilst it can be difficult for researchers to determine what the optimal Sens and Spec values are for their diagnostic test to be considered accurate, the AUC result takes both of these values into account to produce a single value representing the overall diagnostic accuracy of the test, interpreted on an externally validated scale.

Many screening or diagnostic tests use continuous measurement scales, and hence have a score range which permits many possible cut-off values and, therefore, multiple points on a ROC plot, with linear interpolation between the points (the plot tends to a curve as the number of points approaches infinity). For a categorical classifier or predictor with n thresholds, there will be n – 1 points in ROC space. However, for a binary classifier or predictor, there is only one cut-off, a single fixed threshold, and only one potential point, hence a ROC dot rather than a ROC plot. In this circumstance, test accuracy (AUC) is derived from the area of a triangle rather than area under a curve.

When there is only a dichotomous measure, AUC is an accurate index (even if the chosen measure is poor). However, whether ROC plots can be meaningfully applied in the assessment of categorical or continuous measures used as binary classifiers (i.e. dichotomised with a single fixed threshold, as is often the case in clinical practice) is unclear, as it is possible that AUC calculations derived in these circumstances, interpolating between thresholds, may be misleading.^11^ This appears to be seldom recognised in diagnostic accuracy literature. Examples in which ROC plots appear to have been uniformly applied in the assessment of binary classifiers without discussion of whether or not such methodology is valid may be identified in various disciplines.^12-15^

The particular motivation for the current study was therefore to examine the value of ROC plots and AUC calculations when using binary classifiers, compared to categorical and continuous scales. This was done by examining several cognitive screening instruments, although such an evaluation may have broader cross-disciplinary implications for the design of diagnostic accuracy research in which binary predictors are used or considered.^16^

Although some researchers have used ROC plots and AUC calculations to evaluate binary or categorical predictors of cognitive status, including single screening questions,^17^ simple neurological signs,^18^ and categorical decision tree screening instruments,^19^ others have routinely eschewed ROC analysis when examining similar binary or categorical tests.^20-23^

The aims of this study were:

- To construct ROC plots for two clinical signs, the applause sign and the attended with sign, which give a discrete binary classification of cognitive impairment (present/absent), and to calculate AUC using both the rank-sum and DOR methods.
- To construct a ROC plot for Codex, a cognitive screening instrument decision tree with four outcome categories with differing probabilities of dementia, and to calculate and compare Codex AUC values as both a fourfold categorical classifier and as a single fixed threshold binary classifier using both AUC calculation methods.
- To construct a ROC plot for the Mini-Addenbrooke’s Cognitive Examination, a continuous scale cognitive screening instrument, and to calculate and compare AUC values as both a continuous scale and as a single fixed threshold binary classifier using both AUC calculation methods.

## Methods

The datasets of screening test accuracy studies examining the applause sign^20^ and the attended with (AW) sign^23^ were used to construct ROC plots. Both signs provide discrete categorical data (normal/abnormal). In the applause sign, the patient is asked to clap 3 times in imitation of the clinician’s example: clapping 3 times is judged normal and is deemed an indicator of the absence of cognitive impairment, whilst clapping more than 3 times is categorised as abnormal and regarded as an indicator of the presence of cognitive impairment. In the AW sign, attending the cognitive disorders clinic with an accompanying informant is categorised as abnormal, a potential indicator of the presence of cognitive impairment, whilst attending the clinic alone is judged normal (cognitive impairment absent). These two signs were chosen not only because they are binary classifiers but also because one (applause sign) has been reported to be very specific but not very sensitive in screening for cognitive impairment,^20^ whilst the other (AW) is very sensitive but not very specific.^23^

The datasets of screening test accuracy studies examining the cognitive disorders examination (Codex)^19^ and the Mini-Addenbrooke’s Cognitive Examination (MACE)^24^ were also analysed to construct ROC plots. Codex is a two-step decision tree which incorporates components from the Mini-Mental State Examination (three word recall, spatial orientation) along with a simplified clock drawing test to produce four categorical outcomes defining probability of dementia diagnosis (A = very low, B = low, C = high, D = very high). Codex may also be used as a binary classifier by combining categories C and D as a predictor of cognitive impairment^19^ or dementia,^22^ with categories A and B combined as a predictor of the absence cognitive impairment or dementia.

MACE is a cognitive screening instrument widely used in the assessment of patients suspected to have dementia and lesser degrees of cognitive impairment. It comprises tests of attention, memory (7-item name and address), verbal fluency, clock drawing, and memory recall, takes around 5-10 minutes to administer, and has a score range 0-30 (impaired to normal). Previous analyses have established values of AUC for MACE, both by rank-sum and DOR methods.^25,26^ In this study, MACE was analysed both as a continuous ordinal scale and as a binary scale by using a previously defined optimal test cut-off (defined by maximal Youden index) of ≤20/30.^25^

Demographics of the three studies are shown in Table 1. All studies followed the STAndards for the Reporting of Diagnostic accuracy specific for dementia studies.^27^ In all studies subjects gave informed consent and study protocol was approved by the institute’s committee on human research (Walton Centre for Neurology and Neurosurgery Approval: N 310).

**Table 1:** Study demographics

| **Cognitive Screener** | **N** | **Age, median (years)** | **Gender F:M (%F)** | **Prevalence** | **Sens** | **Spec** | **DOR** |
| --- | --- | --- | --- | --- | --- | --- | --- |
| Applause sign | 275 | 61 | 138:137 (50.2) | of dementia = 0.19 | 0.54 | 0.85 | 6.61 |
| Attended with (AW) sign | 1209 | 60 | 588:621 (48.6) | of any cognitive impairment = 0.42 | 0.933 | 0.564 | 18.00 |
| Codex (cut-off: A+B/C+D) | 162 | 61 | 79:83 (49) | of dementia = 0.27 | 0.84 | 0.83 | 25.90 |
| MACE (cut-off: $\leq 20/30$) | 755 | 60 | 352:403 (46.6) | of dementia = 0.15 | 0.912 | 0.707 | 25.06 |

Abbreviations: DOR = diagnostic odds ratio; MACE = Mini-Addenbrooke's Cognitive Examination; Sens = sensitivity; Spec = specificity

For each test, AUC was determined from the ROC plot using the rank-sum method. For a binary classifier, it has been shown^11^ that the value of AUC also simplifies to:

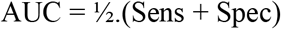

This equation was used to verify the correctness of the AUC value by the rank-sum method, using the values of Sens and Spec extracted from each study dataset (Table 1).

AUC was also determined by calculation from the diagnostic odds ratios (Table 1) using the formula:^10^

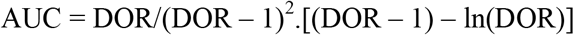

AUC values were classified qualitatively according to the three different schemata.^7-9^

## Results

From the ROC plots constructed for the AW sign (Figure 1) and for the applause sign (not shown), AUC values by the rank-sum method were found to agree in both instances with the calculation from Sens and Spec. AUC calculated from DOR was found to be greater than that calculated by rank-sum method in both cases (Table 2, rows 1 and 2), with consequent changes in the qualitative classification of AUC for both AW sign (in 2/3 schemata) and the applause sign (in 3/3 schemata).

**Table 2:** Cognitive screener AUC values and classification using different methods

| **Cognitive Screener** | **AUC by rank-sum method** | **Classification of AUC calculated by rank-sum method (Metz/Swets/Jones)** | **AUC by DOR method** | **Classification of AUC calculated by DOR method (Metz/Swets/Jones)** |
| --- | --- | --- | --- | --- |
| Applause sign | 0.694 | Poor/low/<good | 0.782 | Fair/moderate/good |
| Attended with (AW) sign | 0.748 | Fair/moderate/<good | 0.879 | Good/moderate/good |
| Codex (fourfold) | 0.856 | Good/moderate/good | 0.904 | Excellent/moderate/good |
| Codex (binary) | 0.836 | Good/moderate/good | 0.904 | Excellent/moderate/good |
| MACE (continuous) | 0.886 | Good/moderate/good | 0.902 | Excellent/moderate/good |
| MACE (binary) | 0.809 | Good/moderate/good | 0.902 | Excellent/moderate/good |

Abbreviations: MACE = Mini-Addenbrooke's Cognitive Examination

**Figure 1:**
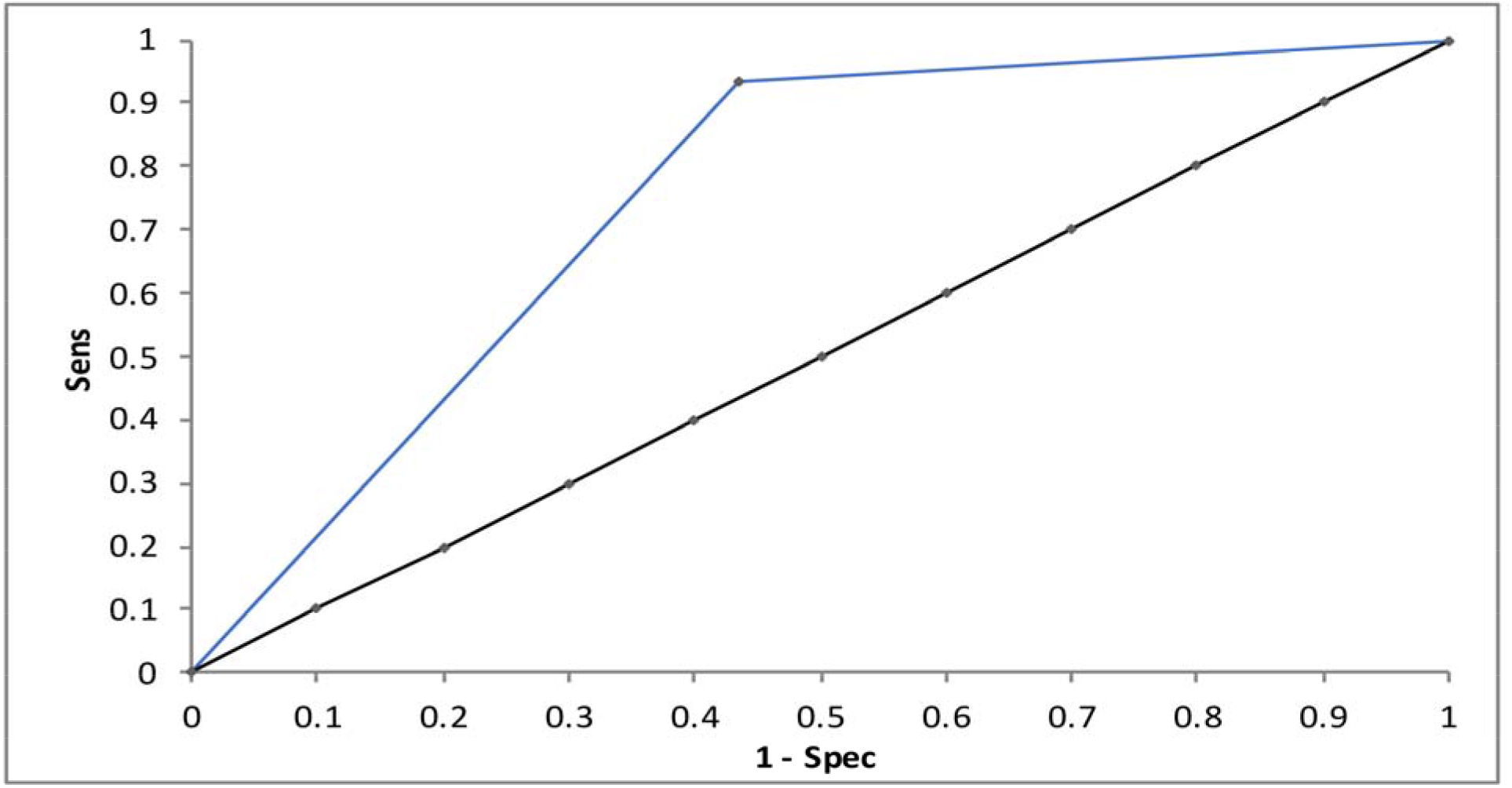
ROC plot for attended with (AW) sign for the diagnosis of any cognitive impairment (dementia + MCI) versus no cognitive impairment, with chance diagonal (y = x)

From the ROC plot constructed for Codex as a fourfold categorical classifier (Figure 2), AUC calculation by DOR method was found to be greater than that by rank-sum method (Table 2, row 3) with some consequent change in qualitative classification of AUC (in 1/3 schemata).

**Figure 2:**
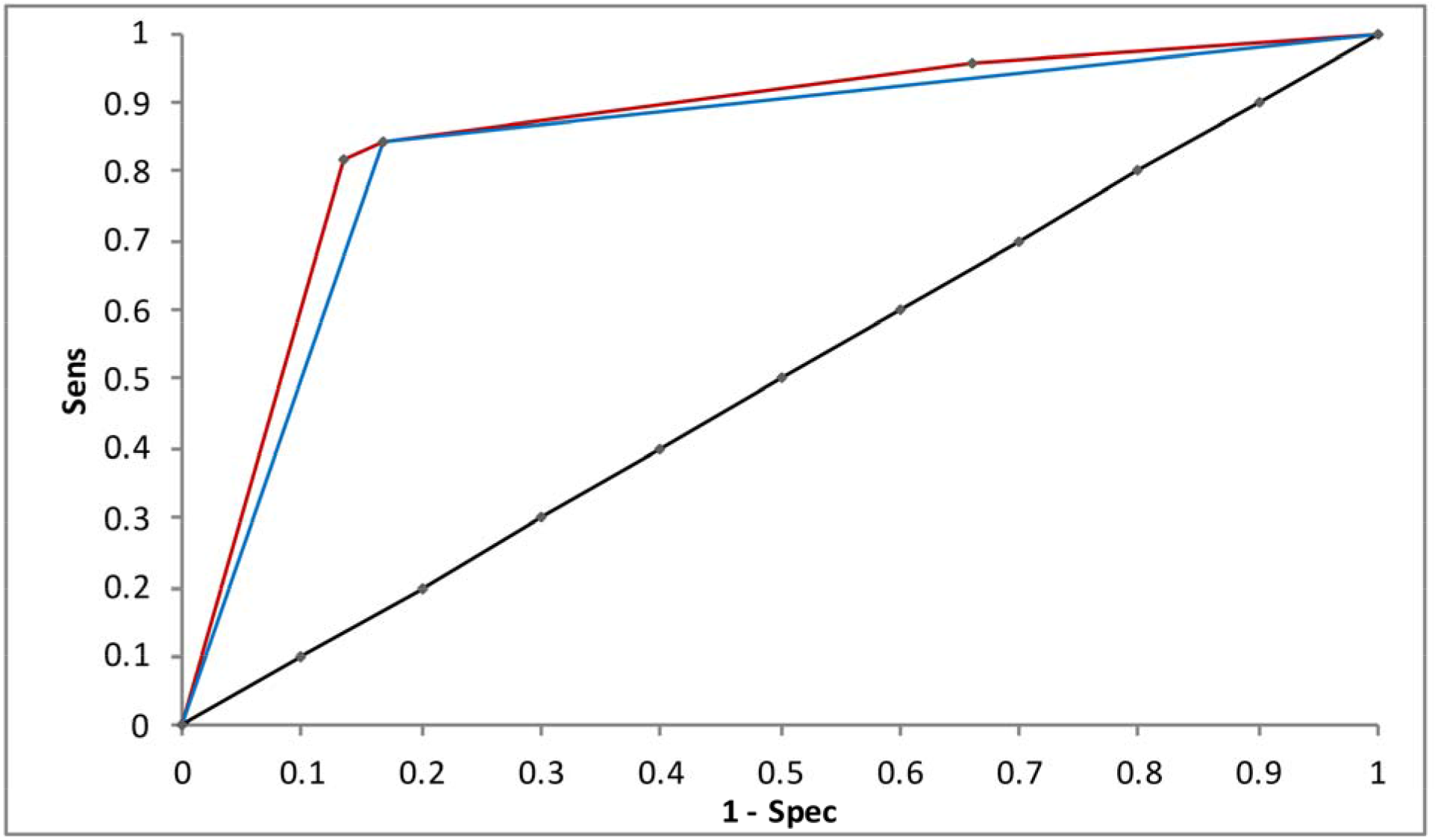
ROC plot for Codex for the diagnosis of dementia versus no dementia, comparing Codex as a fourfold categorical classifier (upper red line) or a binary classifier (lower blue triangle) with chance diagonal (y = x)

From the ROC plot constructed for Codex used as a binary classifier, AUC by rank-sum method was found to agree with the calculation from Sens and Spec. The rank-sum value of AUC was lower for Codex as a binary classifier than as a fourfold categorical classifier (Table 2, rows 3 and 4), the reason for which is apparent when comparing the two ROC plots (Figure 2): the plot as a fourfold classifier lies above the plot as a binary classifier. However, there was no change in the qualitative classifications of AUC. By definition AUC calculated from DOR did not change between Codex used either as a fourfold categorical classifier or as a binary classifier.

From the ROC plot constructed for MACE as a continuous scale (Figure 3), AUC calculated by DOR method was found to be greater than that by rank-sum method (Table 2, row 5), as previously shown,^26^ with some consequent change in qualitative classification of AUC (in 1/3 schemata).

**Figure 3:**
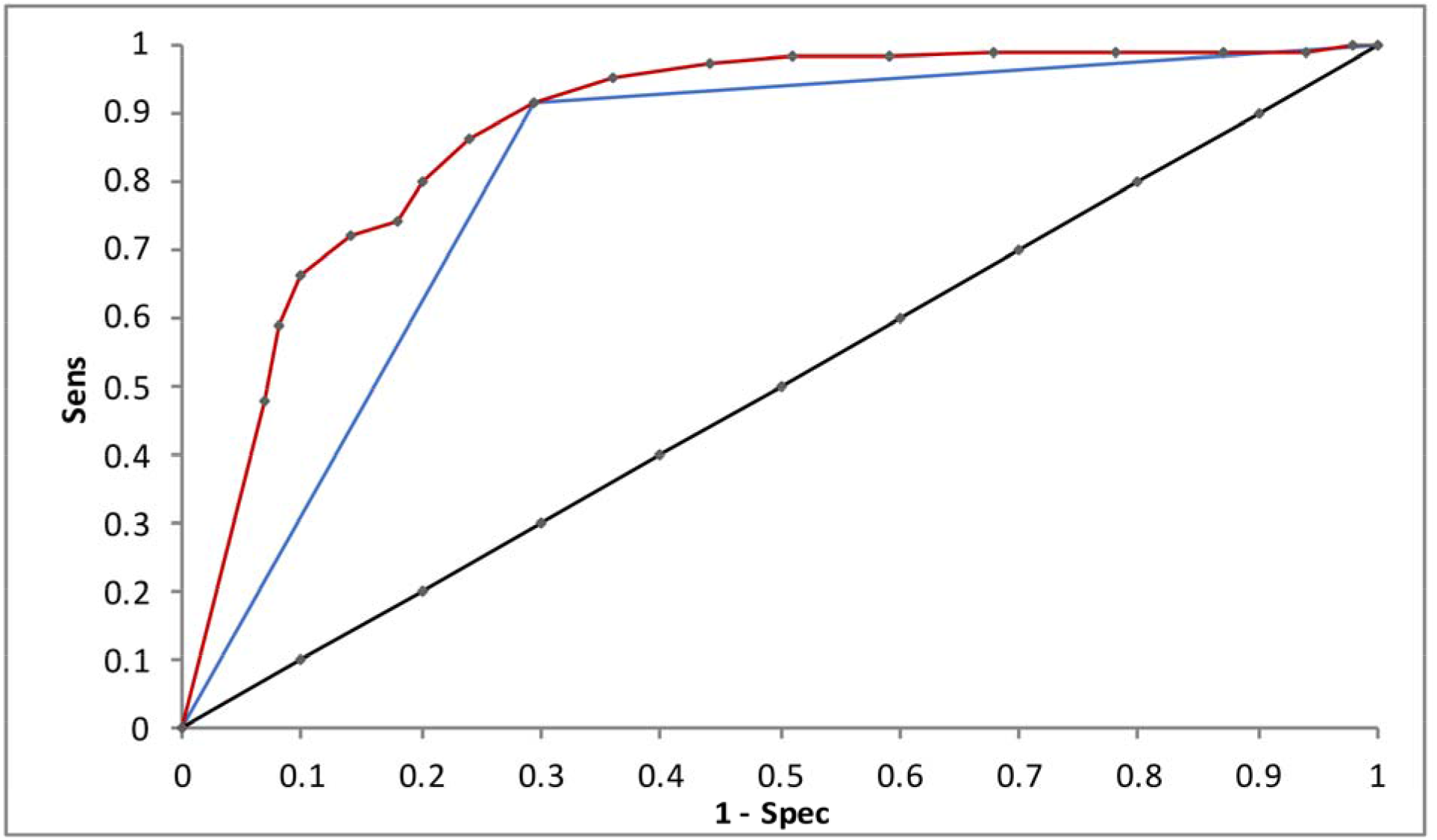
ROC plot for MACE for the diagnosis of dementia versus no dementia, comparing MACE as a continuous scale (upper red curve) or as a binary classifier (lower blue triangle), with chance diagonal (y = x)

From the ROC plot constructed for MACE used as a binary classifier, AUC calculated by rank-sum method was found to agree with the calculation from Sens and Spec. The value of AUC by rank-sum was lower for MACE as a binary classifier compared to MACE as a continuous scale (Table 2, rows 5 and 6), the reason for which is apparent when comparing the two ROC plots (Figure 3) where the plot as a continuous scale lies above the plot as a binary classifier, although there was no change in qualitative classification. By definition AUC calculated from DOR did not change between MACE as either a continuous scale or a binary classifier.

## Discussion

This study has shown that it is possible to apply ROC methodology to the evaluation of studies assessing cognitive screening tests used as binary classifiers, but with certain caveats about the outcomes.

Results from two studies of discrete binary classifiers, examining the applause and AW signs, confirmed that AUC calculated from DOR provided more optimistic values than the usual rank-sum method, as was previously shown for MACE.^26^ The validity of the simplification of AUC calculation for binary categorical data, to the equation AUC = ½.(Sens + Spec),^11^ was also confirmed. The AUC for the AW sign calculated here by rank-sum method (0.75) was inferior to that reported in another study of this sign (0.90).^18^

Results from the studies of cognitive screening instruments which used either a categorical classification (Codex) or a continuous scale (MACE) showed AUC calculated from DOR provided more optimistic values than the usual rank-sum method. AUC values were lower when the tests were used as binary classifiers. These calculations, along with the differences in the graphical representation of the same data when these tests were used as binary predictors (Figures 2 and 3), suggested that the dichotomised measure underrepresented test performance relative to its continuous counterpart. Hence the use of AUC is a potentially misleading metric in these circumstances, as previously suggested.^11^ Although a previous study of Codex included a ROC plot, no AUC was reported.^19^

ROC plots and AUC values are recognised to have various shortcomings in the evaluation of clinical tests. For example, they combine test accuracy over a range of thresholds which may be both clinically relevant and clinically nonsensical,^28^ hence giving an “optimistic” evaluation of test accuracy.^29^ Furthermore, it has been shown that ROC plots and AUC values are unchanged when comparing balanced and imbalanced datasets.^30^ The datasets used in this study were imbalanced with respect to the presence or absence of dementia or cognitive impairment (Table 1, column 5), as is to be anticipated in any clinical population. However, it was not the purpose of this study to compare ROC and AUC performance in balanced versus unbalanced datasets, but to examine “real world” clinical data.

Clinicians (and indeed patients) may generally be said to prefer binary classifiers (e.g. screen positive vs screen negative; target diagnosis present vs absent) since they give an impression of certainty. Indeed, one of the reasons for undertaking ROC analysis of test accuracy study data is to define optimal test cut-offs, for example using maximal Youden index or minimal Euclidean index,^31^ also known as dichotomisation points or decision thresholds, so that tests generating continuous scale data may be used as if they were binary classifiers.^3^ Whilst this may prove useful at a clinical level, there are potential penalties for dichotomising a continuous variable such as cognitive function, as greater statistical power is afforded by the continuous approach.^32^ Hence previous diffidence in applying ROC methodology to the assessment of clinical signs and tests providing binary or categorical data^20-23^ may not have been unjustified.

More generally, the findings from this study may have implications beyond the use of cognitive screening instruments. In situations where the continuous scale approach is unavailable, for example in studies assessing inherently binary outcomes such as mortality or the diagnostic accuracy of administrative healthcare data,^16^ it may be challenging for researchers to demonstrate the relative trade-off between test benefits and costs beyond taking a narrative (subjective) approach to conveying the magnitude of these differences. In such restricted circumstances, an approach with less bias than the narrative one may be to apply ROC methodology to the binary classifier, but ensuring that the limitations are made clear. This approach should be used only for internal assessment of the relative differences, in terms of costs-benefits trade-off, between different within-study case ascertainment algorithms, rather than for any external comparison of the results against other datasets or cut-offs for “interpretation”. This is because, as shown, comparison of binary ROC methodology against external standards or continuous methods may be misleading. Future research will be required to indicate whether there are any standardised ways to interpret ROC results for binary classifiers, in a way that may have external study validity, given the availability and potential necessity of such an approach in certain rare circumstances.

In conclusion, this study has demonstrated that ROC plots and AUC values using categorical or continuous scale tests as binary classifiers may be misleading for the purposes of clinical decision making. Various statistical packages will readily allow researchers to calculate AUC values with a binary classifier,^33-35^ and therefore this study may help researchers interpret the results of such analysis within the context of their potential limitations. In circumstances where this is deemed the best available analysis method, researchers should be aware of the limitations and make them clear in their work.

## Data Availability

Available on reasonable request

## Notes

### Competing Interest Statement

The authors have declared no competing interest.

### Funding Statement

None

### Author Declarations

Walton Centre for Neurology and Neurosurgery

